# Telomere Maintenance Is Acquired at Liposarcoma Dedifferentiation, and Genetically Longer Telomeres Are Associated With Liposarcoma Risk

**DOI:** 10.64898/2026.09.28.26364135

**Authors:** Minggui Pan, Joyce Chi, Lillian Zhi, Benjamin Lin, Maggie Yuxi Zhou, Nam Bui, Kristen Ganjoo

## Abstract

The contribution of telomere length, telomerase activation, and alternative lengthening of telomeres (ALT) to individual sarcoma subtypes remains unclear. We analyzed 11,792 patients with sarcoma sequenced across 19 centers in AACR Project GENIE v20.0, defining telomerase activation as *TERT* promoter mutation, amplification, or rearrangement and using ATRX/DAXX inactivation as the ALT proxy, with denominators restricted to patients whose assay interrogated the relevant locus. Findings were tested for *TERT* expression in The Cancer Genome Atlas, for overall survival in MSK-IMPACT 50K, and for replication in an independent Foundation Medicine cohort; Mendelian randomization in FinnGen tested germline telomere length. Telomerase activation (8.8%) and ATRX/DAXX loss (8.0%) segregated by histology and co-occurred less often than expected across subtypes (Mantel-Haenszel odds ratio [OR], 0.52; 95% CI, 0.29 to 0.92). In *MDM2*-amplified dedifferentiated liposarcoma (DDLPS), telomerase was activated through *TERT* amplification (17.2%) or rearrangement (2.7%) and never through promoter mutation (0/261), and never co-occurred with ATRX/DAXX loss (0/49; *P* = .011). *TERT*-amplified tumors expressed *TERT* (*P* = .002), and amplification was largely focal. Telomere maintenance alterations were present in 28.0% of DDLPS versus 8.7% of well-differentiated liposarcoma (*P* < .001) and predicted shorter overall survival in DDLPS (hazard ratio, 2.04; 95% CI, 1.23 to 3.39; *P* = .005). ATRX/DAXX loss was more frequent in uterine than extrauterine leiomyosarcoma in GENIE (adjusted OR, 1.96) and in the independent cohort (OR, 1.95; both *P* < .001). Genetically longer telomeres were associated with liposarcoma (OR per SD, 4.10; 95% CI, 2.15 to 7.81). Telomere maintenance in sarcoma is therefore histology-specific; in liposarcoma it is acquired at dedifferentiation through focal *TERT* amplification or ALT rather than promoter mutation, and it identifies DDLPS with inferior survival.

## INTRODUCTION

Telomerase reactivation is common in carcinomas and sarcomas, frequently through hotspot mutations in the *TERT* promoter that create de novo ETS-binding sites.^1,2^ Some tumors instead use alternative lengthening of telomeres (ALT), a homologous recombination-based mechanism tightly linked to inactivation of the chromatin remodelers ATRX or DAXX.^3,4^ Tumors of mesenchymal origin (sarcomas) use ALT at a markedly higher frequency than epithelial cancers (carcinomas).^5,6^ Germline defects in telomere and mitotic function also selectively predispose to sarcoma,^7^ placing telomere biology near the center of sarcomagenesis.

Individual sarcoma subtypes favor one mechanism or the other: *TERT* promoter mutations are recurrent in myxoid liposarcoma and solitary fibrous tumor,^8–10^ whereas ALT and ATRX loss are frequent in leiomyosarcoma, pleomorphic liposarcoma, and dedifferentiated liposarcoma (DDLPS).^5,6,11^ The single-institution studies underlying these observations were limited by cohort size, by reliance on promoter sequencing without copy-number assessment, and by the absence of cross-subtype comparison under a uniform analytic framework.

We asked whether telomerase activation in sarcoma is confined to promoter mutation or also proceeds through *TERT* amplification and rearrangement, and whether the two mechanisms are mutually exclusive across the histologic spectrum. We then asked when telomere maintenance is acquired during progression, comparing well-differentiated liposarcoma (WDLPS) with DDLPS. Both are defined by 12q13-15 amplification of *MDM2* and *CDK4*, and DDLPS arises from WDLPS through progression to a nonlipogenic, higher-grade component with a markedly worse prognosis.^12,13^

Whether acquiring a telomere maintenance mechanism accompanies this transition has not been examined at scale.

We used AACR Project GENIE, a multi-institutional registry of clinical-grade tumor sequencing,^14^ to characterize telomere maintenance alterations in 11,792 patients with sarcoma.

## MATERIALS AND METHODS

### Data Source and Cohort

We accessed AACR Project GENIE v20.0-public (released July 2026) through the GENIE cBioPortal application programming interface.^14,15^ Samples with Cancer Type annotated as soft tissue sarcoma, bone cancer, uterine sarcoma, gastrointestinal stromal tumor, breast sarcoma, undifferentiated embryonal sarcoma of the liver, clear cell sarcoma of kidney, or infantile fibrosarcoma, together with malignant peripheral nerve sheath tumor, were eligible (14,443 samples; 12,919 patients). The tumors of benign, intermediate, and non-mesenchymal entities (including hemangioma, desmoid fibromatosis, tenosynovial giant cell tumor, giant cell tumor of bone, myxoma, uterine leiomyoma and smooth muscle tumor of uncertain malignant potential, benign and borderline phyllodes tumor, inflammatory myofibroblastic tumor, and dendritic cell sarcomas) were excluded. One sample per patient was retained, prioritizing primary tumors and then the assay with the broadest coverage of the loci studied, yielding 11,792 patients.

We grouped OncoTree codes into histologic entities: all leiomyosarcoma codes (soft tissue, uterine, myxoid, and epithelioid) as leiomyosarcoma, with uterine origin recorded as a covariate; malignant fibrous histiocytoma with undifferentiated pleomorphic sarcoma (UPS); osteosarcoma variants; skeletal and extraskeletal Ewing sarcoma; chordoma variants; and atypical lipomatous tumor with WDLPS. Not-otherwise-specified codes contributed to pooled estimates but were not reported as subtypes. Liposarcoma analyses were restricted to tumors with *MDM2* amplification unless stated otherwise.

### Definition of Telomere Maintenance Alterations

*TERT* promoter mutation was defined as any variant in chr5:1,295,141-1,295,315 (GRCh37), encompassing the C228T, C250T, and C242/243 hotspots. *TERT* amplification was defined as a high-level gain (GISTIC-like discrete value of 2) and *TERT* rearrangement as any structural variant involving *TERT*. ATRX or DAXX inactivation was defined as a truncating variant (nonsense, frameshift, canonical splice site, translation start site, or nonstop), homozygous deletion, or structural variant; missense variants were excluded because most are of uncertain significance.

### Assay-Aware Evaluability

GENIE aggregates 88 sequencing assays in this cohort, which differ in gene content and in capture of the *TERT* promoter. A patient contributed to the denominator for an alteration only if the assay interrogated that locus. Because cBioPortal gene panel definitions list *TERT* at the gene level without distinguishing promoter capture, we defined promoter evaluability empirically: an assay was considered promoter-evaluable only if it had reported at least three *TERT* promoter variants across all 289,869 GENIE samples. This excluded several assays that list *TERT* but contribute no promoter calls, including MSK-IMPACT505 (0 promoter calls among 45,718 samples). Copy number-based alterations were counted only in samples profiled for copy number.

### Validation Cohorts

Three cBioPortal cohorts were used for validation. TCGA-SARC (n = 206), which is independent of GENIE, provided RNA sequencing (RSEM) for *TERT* expression and GISTIC copy number.^24^ MSK-IMPACT 50K (54,331 tumors; 4,154 sarcoma samples) provided overall survival measured from the date of sequencing, arm-level copy number, and genetic ancestry;^25^ because these patients overlap the MSK contribution to GENIE, this cohort was used for outcome analyses rather than prevalence replication. Diagnosis and sequencing dates for a subset of patients were obtained from MSK-MET timelines.^26^ An independent Foundation Medicine cohort of 7,494 sarcomas, profiled for mutations and structural variants but not copy number, was used to replicate the leiomyosarcoma site association.^27^ Alteration definitions matched the primary analysis within the limits of each platform.

### Mendelian Randomization

To test whether germline telomere length influences sarcoma susceptibility, we performed two-sample MR with public summary statistics. Outcomes were the sarcoma endpoints of FinnGen release 12 with at least 100 cases, as defined by FinnGen: liposarcoma (328 cases), leiomyosarcoma (306), gastrointestinal stromal tumor and stromal sarcoma (351), fibrosarcoma (280), chondrosarcoma (158), osteosarcoma (132), and malignant neoplasm of bone and articular cartilage (648), each compared with controls without any cancer diagnosis.^28^ Instruments for leukocyte telomere length were the 124 conditionally independent variants designated for MR in a UK Biobank genome-wide association study of 472,174 participants, with European-ancestry effects expressed per SD.^29^ Height and body mass index, anthropometric traits associated with the risk of several cancers, served as comparator exposures, using variants reaching *P* < 5 × 10^−8^ in the GIANT-UK Biobank meta-analysis of approximately 700,000 individuals, pruned to the strongest variant per 1-Mb window.^30^ Variants were mapped to GRCh38 and aligned to FinnGen alleles; palindromic variants were retained only when the minor allele frequency was 0.42 or less and were oriented by allele frequency.

The primary estimator was multiplicative random-effects inverse-variance weighting (IVW).^31^ Sensitivity analyses comprised MR-Egger regression,^32^ the weighted median,^33^ leave-one-out analysis, and exclusion of palindromic variants. For telomere length, a further analysis excluded variants at canonical telomere-biology loci, including *TERT*, *TERC*, and *PARP1*, to limit pleiotropy acting through telomerase itself. The false discovery rate was controlled across the 21 exposure-outcome tests, and the minimum OR detectable at 80% power was derived from each IVW standard error. UK Biobank and FinnGen do not share participants, so bias from sample overlap is not expected.

### Statistical Analysis

Prevalence estimates are reported with Wilson 95% CIs. Mutual exclusivity was assessed within subtypes with Fisher exact tests and across subtypes with the Mantel-Haenszel common odds ratio (OR) and the Breslow-Day test of homogeneity, using subtypes with at least 30 dual-evaluable patients. Associations with age (per 10 years), sex, race, and anatomic site were estimated with Firth-penalized logistic regression,^16,17^ which remains unbiased under sparse events, adjusting for contributing center (centers with fewer than 20 patients pooled). Heterogeneity of age effects across subtypes was tested with a penalized likelihood ratio test for the subtype-by-age interaction. The Benjamini-Hochberg procedure controlled the false discovery rate across subtypes. All tests were two-sided. Overall survival was analyzed with Cox proportional hazards models adjusted for age, sample type, and metastatic disease status at sequencing; sensitivity analyses restricted to primary tumors and measured survival from diagnosis with delayed entry at sequencing to account for left truncation. Analyses used Python 3.11 with pandas, SciPy, and statsmodels.

## RESULTS

### Characteristics of the cohorts

The analytic cohort comprised 11,792 patients from 19 contributing centers (Table 1). Median age at sequencing was 56 years (IQR, 38-67); 9.5% were younger than 18 years. Soft tissue sarcoma accounted for 56.4% of patients, gastrointestinal stromal tumor for 18.3%, bone sarcoma for 13.1%, and uterine sarcoma for 8.4%. ATRX and DAXX were evaluable in 9,766 patients (82.8%), the *TERT* promoter in 6,233 (52.9%), and all three *TERT* mechanisms in 4,891 (41.5%).

**Table 1.** Patient and Sample Characteristics (N = 11,792)

| Characteristic | No. (%) |
| --- | --- |
| Patients | 11,792 |
| Contributing centers | 19 |
| Sequencing assays | 88 |
| Age at sequencing, years, median (IQR) | 56 (38-67) |
| Age <18 years | 1,118 (9.5) |
| Sex, female | 5,757 (48.8) |
| Sex, male | 4,978 (42.2) |
| Sex, unknown | 1,057 (9.0) |
| Race, white | 6,975 (59.2) |
| Race, black | 828 (7.0) |
| Race, asian | 712 (6.0) |
| Race, other/unknown | 3,277 (27.8) |
| Sample, primary | 7,542 (64.0) |
| Sample, metastasis | 3,112 (26.4) |
| Sample, other/not collected | 1,138 (9.7) |
| Cancer type, Soft Tissue Sarcoma | 6,654 (56.4) |
| Cancer type, Gastrointestinal Stromal Tumor | 2,153 (18.3) |
| Cancer type, Bone Cancer | 1,548 (13.1) |
| Cancer type, Uterine Sarcoma | 996 (8.4) |
| Cancer type, Nerve Sheath Tumor | 251 (2.1) |
| Cancer type, Breast Sarcoma | 163 (1.4) |
| Cancer type, Undifferentiated Embryonal Sarcoma of the Liver | 12 (0.1) |
| Cancer type, Clear Cell Sarcoma of Kidney | 9 (0.1) |
| Cancer type, Infantile Fibrosarcoma | 6 (0.1) |
| Evaluable, TERT promoter | 6,233 (52.9) |
| Evaluable, TERT promoter + copy number | 4,891 (41.5) |
| Evaluable, ATRX and DAXX | 9,766 (82.8) |

NOTE. One sample per patient. Evaluability denotes that the sequencing assay interrogated the locus (see Methods).

### Telomere Maintenance Mechanisms Segregate by Histology

Telomerase activation was identified in 8.8% (432/4,891) and ATRX/DAXX loss in 8.0% (785/9,766) of evaluable patients (Table 2). Promoter mutations involved C228T in 292 of 349 tumors (83.7%) and C250T in 45 (12.9%). ATRX accounted for nearly all ALT-proxy events (794 ATRX *v* 43 DAXX inactivations in their respective evaluable populations).

**Table 2.** Telomere Maintenance Alterations by Sarcoma Subtype.

| Subtype | Patients | *TERT* promoter mutation | *TERT* amplification | *TERT* activation (any) | ATRX/DAXX loss |
| --- | --- | --- | --- | --- | --- |
| Myxoid/round cell liposarcoma | 174 | 77/97 (79.4) | 0/78 (0.0) | 64/78 (82.1) | 0/162 (0.0) |
| Solitary fibrous tumor | 239 | 57/126 (45.2) | 0/103 (0.0) | 45/103 (43.7) | 0/203 (0.0) |
| Malignant phyllodes tumor | 60 | 22/45 (48.9) | 0/39 (0.0) | 17/39 (43.6) | 0/55 (0.0) |
| Dedifferentiated liposarcoma | 581 | 1/295 (0.3) | 46/269 (17.1) | 51/269 (19.0) | 55/534 (10.3) |
| Clear cell sarcoma | 71 | 8/40 (20.0) | 0/30 (0.0) | 5/30 (16.7) | 0/55 (0.0) |
| MPNST | 251 | 6/167 (3.6) | 10/120 (8.3) | 15/120 (12.5) | 8/220 (3.6) |
| Undifferentiated pleomorphic sarcoma | 652 | 31/389 (8.0) | 8/273 (2.9) | 33/273 (12.1) | 106/573 (18.5) |
| Chondrosarcoma | 259 | 10/140 (7.1) | 2/95 (2.1) | 9/95 (9.5) | 5/205 (2.4) |
| Uterine adenosarcoma | 106 | 1/49 (2.0) | 2/37 (5.4) | 3/37 (8.1) | 8/89 (9.0) |

| Subtype | Patients | TERT promoter mutation | TERT amplification | TERT activation (any) | ATRX/DAXX loss |
| --- | --- | --- | --- | --- | --- |
| Osteosarcoma | 613 | 11/389 (2.8) | 13/276 (4.7) | 20/276 (7.2) | 51/516 (9.9) |
| Well-differentiated liposarcoma | 246 | 2/82 (2.4) | 3/71 (4.2) | 5/71 (7.0) | 2/218 (0.9) |
| DSRCT | 178 | 6/88 (6.8) | 0/80 (0.0) | 5/80 (6.2) | 1/170 (0.6) |
| Embryonal rhabdomyosarcoma | 164 | 2/98 (2.0) | 2/82 (2.4) | 5/82 (6.1) | 4/146 (2.7) |
| PEComa | 121 | 3/78 (3.8) | 1/66 (1.5) | 4/66 (6.1) | 21/110 (19.1) |
| Chordoma | 178 | 5/105 (4.8) | 0/63 (0.0) | 3/63 (4.8) | 6/166 (3.6) |
| Rhabdomyosarcoma, other | 253 | 2/154 (1.3) | 3/100 (3.0) | 4/100 (4.0) | 9/223 (4.0) |
| Ewing sarcoma | 453 | 8/282 (2.8) | 0/194 (0.0) | 6/194 (3.1) | 1/367 (0.3) |
| Leiomyosarcoma | 1558 | 15/780 (1.9) | 8/644 (1.2) | 18/644 (2.8) | 300/1281 (23.4) |
| Angiosarcoma | 397 | 8/231 (3.5) | 0/194 (0.0) | 5/194 (2.6) | 16/353 (4.5) |
| GIST | 2153 | 6/1011 (0.6) | 14/826 (1.7) | 19/826 (2.3) | 2/1441 (0.1) |
| Pleomorphic liposarcoma | 90 | 1/52 (1.9) | 0/47 (0.0) | 1/47 (2.1) | 19/80 (23.8) |
| Synovial sarcoma | 298 | 0/179 (0.0) | 2/147 (1.4) | 2/147 (1.4) | 3/260 (1.2) |
| Myxofibrosarcoma | 255 | 0/111 (0.0) | 1/90 (1.1) | 1/90 (1.1) | 45/229 (19.7) |
| Epithelioid hemangioendothelioma | 101 | 0/62 (0.0) | 0/47 (0.0) | 0/47 (0.0) | 4/87 (4.6) |
| Epithelioid sarcoma | 94 | 0/52 (0.0) | 0/43 (0.0) | 0/43 (0.0) | 1/82 (1.2) |
| Alveolar rhabdomyosarcoma | 115 | 0/70 (0.0) | 0/41 (0.0) | 0/41 (0.0) | 0/98 (0.0) |
| All sarcomas | 11,792 | 349/6233 (5.6) | 151/4891 (3.1) | 432/4891 (8.8) | 785/9766 (8.0) |

NOTE. Values are altered/evaluable (%). Denominators differ by alteration because assays differ in locus coverage; TERT activation (any) requires both promoter and copy-number evaluability. Subtypes with at least 40 ATRX/DAXX-evaluable patients are shown; liposarcoma rows are not restricted by MDM2 status (MDM2-restricted estimates are given in the text). NE, not evaluable.
Abbreviations: DSRCT, desmoplastic small round cell tumor; GIST, gastrointestinal stromal tumor; MPNST, malignant peripheral nerve sheath tumor; PEComa, perivascular epithelioid cell tumor.

Subtypes separated into three groups (Fig 1A). A telomerase-dominant group relied almost exclusively on *TERT* promoter mutation: myxoid/round cell liposarcoma (79.4%), malignant phyllodes tumor (48.9%), solitary fibrous tumor (45.2%), and clear cell sarcoma (20.0%), none with ATRX/DAXX loss. An ALT-dominant group comprised pleomorphic liposarcoma (23.8%), leiomyosarcoma (23.4%), myxofibrosarcoma (19.7%), PEComa (19.1%), and UPS (18.5%). A third group of translocation-driven sarcomas, including Ewing sarcoma, synovial sarcoma, alveolar rhabdomyosarcoma, desmoplastic small round cell tumor, and epithelioid sarcoma, had few or no alterations in either pathway.

**Fig 1.**
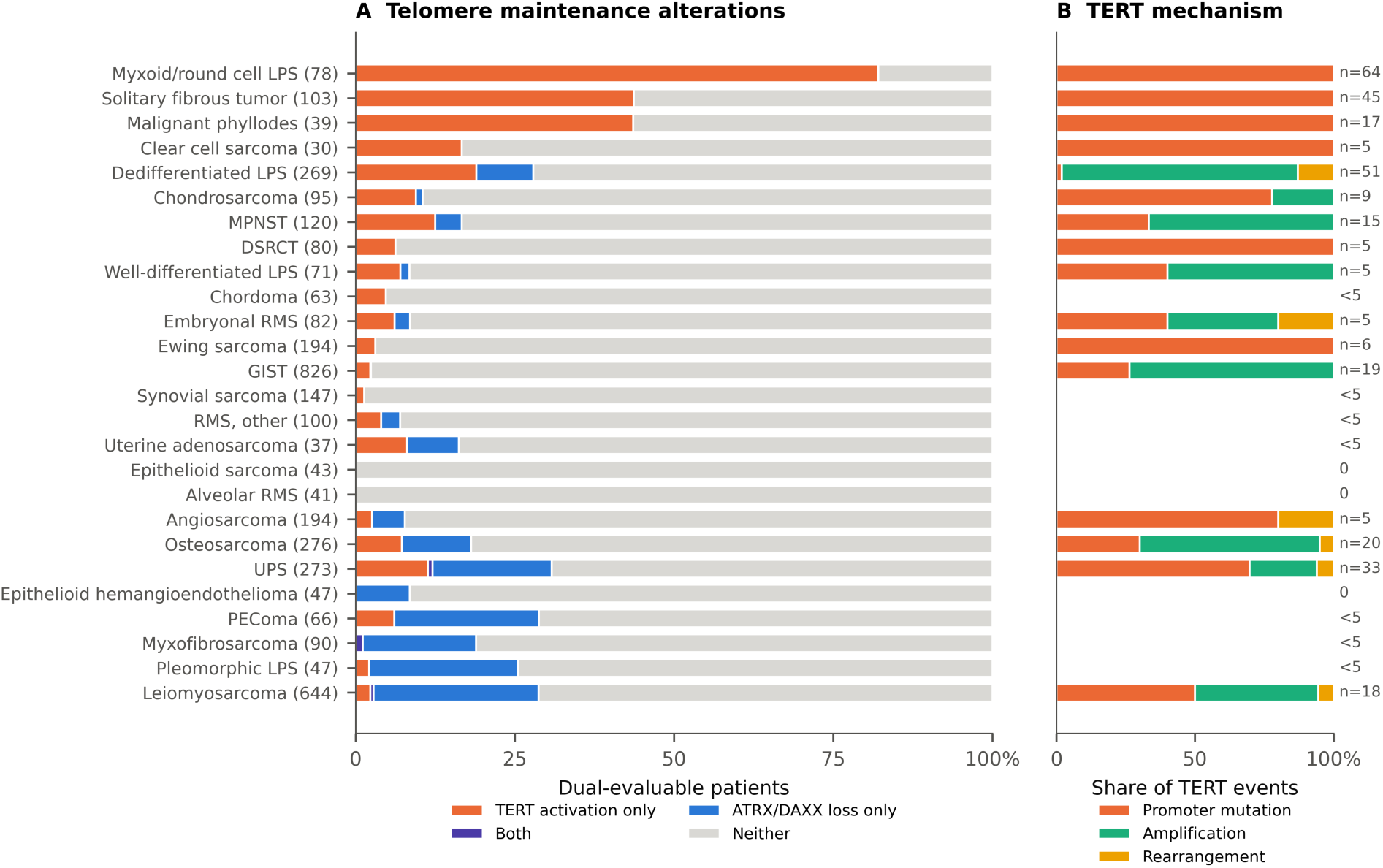
Telomere maintenance mechanisms across sarcoma subtypes. (A) Proportion of dual-evaluable patients with telomerase activation only (TERT promoter mutation, amplification, or rearrangement), ATRX/DAXX loss only, both, or neither; subtypes with at least 30 dual-evaluable patients, ordered by net telomerase-versus-ALT balance. Patient numbers in parentheses. (B) Mechanism of TERT* activation among telomerase-activated tumors; shown for subtypes with at least five events

The mechanism of telomerase activation also differed by histology (Fig 1B). Promoter mutation accounted for all *TERT* events in myxoid/round cell liposarcoma, solitary fibrous tumor, and malignant phyllodes tumor. In contrast, *TERT* amplification is the predominant alteration in DDLPS, osteosarcoma, malignant peripheral nerve sheath tumor, and gastrointestinal stromal tumor.

### Telomerase Activation and ATRX/DAXX Loss Are Mutually Exclusive

Across 15 subtypes with sufficient events, telomerase activation and ATRX/DAXX loss co-occurred less often than expected (Mantel-Haenszel OR, 0.52; 95% CI, 0.29 to 0.92; *P* = .019), with the degree of exclusivity varying by histology (Breslow-Day *P* = .012) and was strongest in DDLPS and UPS. When telomerase activation was restricted to promoter mutation, the pooled association was attenuated and no longer significant (OR, 0.76; 95% CI, 0.40 to 1.43; *P* = .37), indicating that amplification and rearrangement exerted most of the impact of the exclusivity signal.

### Liposarcoma: Telomere Maintenance Is Acquired at Dedifferentiation

*MDM2* amplification was detected in 96.1% of copy number-profiled DDLPS and 86.8% of WDLPS, confirming the diagnostic assignment. Among *MDM2*-amplified DDLPS, telomerase activation occurred in 18.8% (49/261; 95% CI, 14.5 to 24.0) and ATRX/DAXX loss in 9.6% (46/478; 95% CI, 7.3 to 12.6). In myxoid/round cell liposarcoma, *MDM2*-amplified DDLPS was found to carry zero *TERT* promoter mutation (0/261; 95% CI, 0 to 1.5), 17.2% *TERT* amplification and 2.7% *TERT* rearrangement (Fig 2A). *TERT* amplification was observed on both MSK-IMPACT (31/189) and DFCI-OncoPanel (14/69) assays.

**Fig 2.**
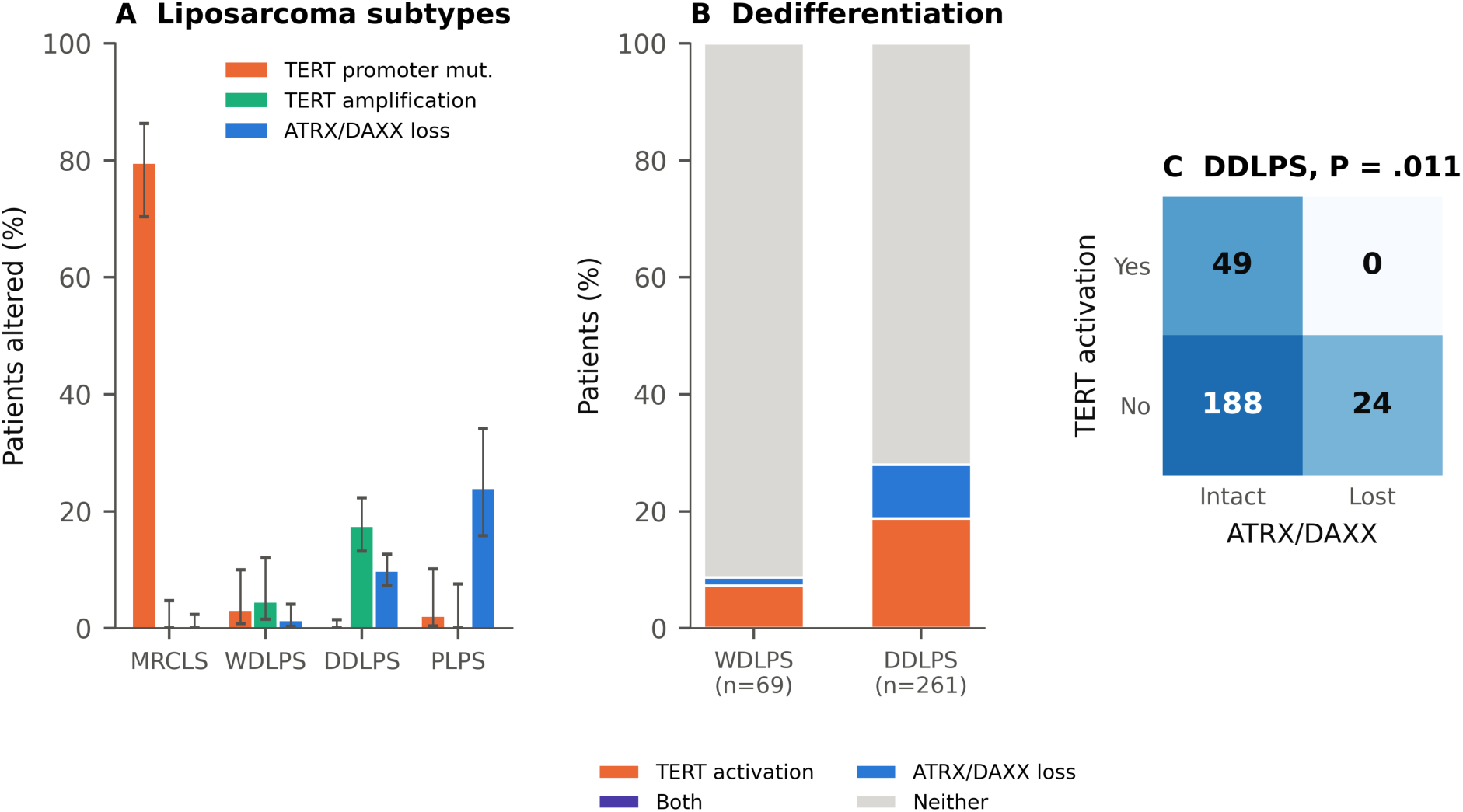
Telomere maintenance in liposarcoma. (A) Prevalence of TERT promoter mutation, TERT amplification, and ATRX/DAXX loss in myxoid/round cell liposarcoma (MRCLS), MDM2-amplified well-differentiated (WDLPS) and dedifferentiated liposarcoma (DDLPS), and pleomorphic liposarcoma (PLPS). Error bars, Wilson 95% CIs. (B) Composition of telomere maintenance status in MDM2-amplified WDLPS and DDLPS among dual-evaluable patients (P < .001). (C) Mutual exclusivity of telomerase activation and ATRX/DAXX loss in DDLPS (Fisher exact P* = .011)

None of 49 telomerase-activated tumors had ATRX/DAXX loss, compared with 24 of 212 without telomerase activation (*P* = .011; Fig 2C). *TERT* amplification rates did not differ by *CDK4* amplification (17.3% *v* 13.3%) or *JUN* amplification (23.1% *v* 14.5%; *P* = .26). Telomerase activation increased with age in DDLPS (OR per 10 years, 1.40; 95% CI, 1.06 to 1.87; *P* = .019), whereas ATRX/DAXX loss did not increase (OR, 0.87; *P* = .28). Among the 21 tumors annotated as DDLPS without *MDM2* amplification, ATRX/DAXX loss was more frequent (5/17, 29.4% *v* 9.6%; *P* = .023), consistent with misclassified pleomorphic liposarcoma or UPS.

Alteration in telomere maintenance pathway was strongly enriched at dedifferentiation (Fig 2B), present in 28.0% of *MDM2*-amplified DDLPS (73/261; 95% CI, 22.9 to 33.7) compared with 8.7% of WDLPS (6/69; 95% CI, 4.0 to 17.7; *P* < .001). After adjustment for age, sex, and center, DDLPS was associated with higher odds of *TERT* amplification (OR, 3.57; 95% CI, 1.16 to 11.03; *P* = .027) and of ATRX/DAXX loss (OR, 7.17; 95% CI, 2.02 to 25.41; *P* = .002).

### Validation

In TCGA-SARC, *TERT*-amplified sarcomas expressed *TERT* (median log2 RSEM, 4.64 *v* 0.00 in tumors with neither alteration; *P* = .002; Fig 4A). Within DDLPS, 6 of 50 tumors were *TERT*-amplified, 9 had ATRX/DAXX loss, and 1 had both (*P* = .016). This pattern was reproduced by the MSK-IMPACT 50K cohort. In *MDM2*-amplified DDLPS (n = 226), *TERT* amplification was found in 34 (15.0%), rearrangement in 7, promoter mutation in 1, ATRX/DAXX loss in 21 (9.3%), and no tumor with both alterations. Among *TERT*-amplified DDLPS with arm-level calls, the 5p arm was unchanged in 9 of 17 (53%), indicating a focal 5p15.33 amplicon rather than arm-level gain (Fig 4B).

Among 208 patients with *MDM2*-amplified DDLPS and survival data (80 deaths), median OS was 22.1 months with a telomere maintenance alteration versus 54.9 months without (log-rank *P* = .008; Fig 4C). After adjustment for age, sample type, and metastatic disease at sequencing, the hazard ratio (HR) was 2.04 (95% CI, 1.23 to 3.39; *P* = .005). The association was primarily driven by telomerase activation (HR, 2.05; 95% CI, 1.14 to 3.67; *P* = .016), but not ATRX/DAXX loss (HR, 1.61; 95% CI, 0.76 to 3.41; *P* = .22), This was consistently identified when analyzing the primary tumors only (HR, 2.06; 95% CI, 1.11 to 3.83; *P* = .022) and when survival was measured from diagnosis with delayed entry at NGS (n = 127; HR, 1.96; 95% CI, 1.08 to 3.55; *P* = .027). In the 50 case of TCGA DDLPS, the estimate had a large confidence interval (HR, 1.04; 95% CI, 0.42 to 2.55).

### ALT in Leiomyosarcoma

Among 1,558 patients with leiomyosarcoma, ATRX/DAXX loss was more frequent in uterine than extrauterine primaries (31.8% [135/425] *v* 19.3% [165/856]; adjusted OR, 1.96; 95% CI, 1.49 to 2.58; *P* < .001; Fig 3), whereas telomerase activation was uncommon at both sites (3.9% *v* 2.1%; *P* = .34). An apparent association of female sex with ATRX/DAXX loss in the pooled leiomyosarcoma model reflected uterine origin; within extrauterine leiomyosarcoma, sex was not associated with ATRX/DAXX loss (OR for male sex, 0.71; 95% CI, 0.46 to 1.09; *P* = .12). This site association was replicated in the independent Foundation Medicine cohort, in which ATRX/DAXX inactivation was present in 23.8% of uterine (129/541) versus 13.9% of soft tissue leiomyosarcomas (132/952; OR, 1.95; *P* < .001), an effect size nearly identical to that in GENIE.

**Fig 3.**
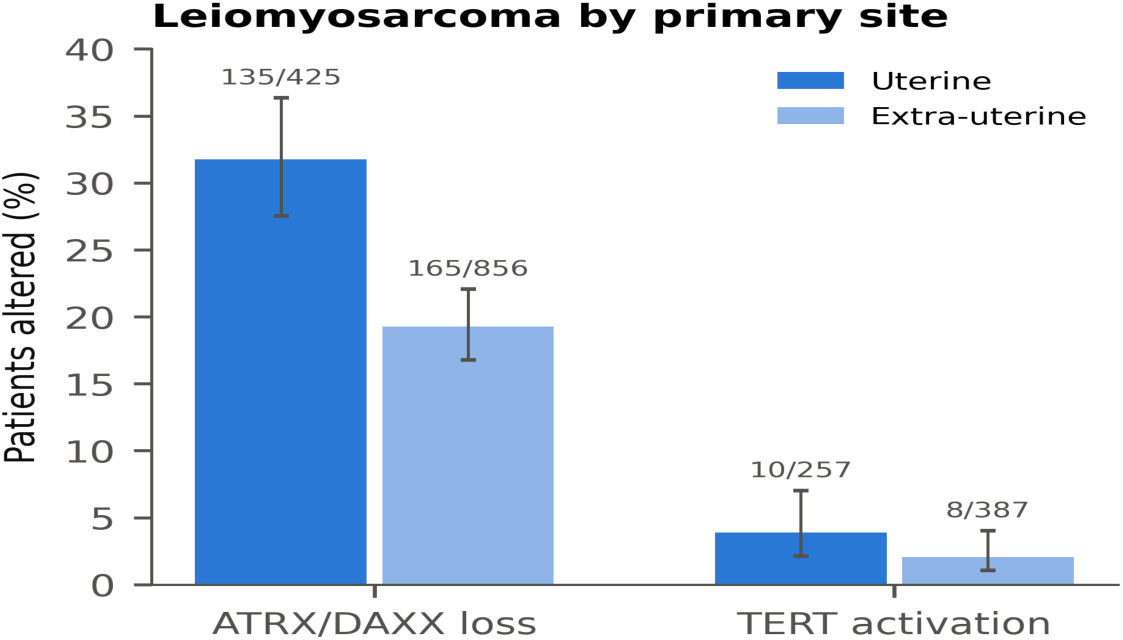
ATRX/DAXX loss and telomerase activation in leiomyosarcoma by primary site. Error bars, Wilson 95% CIs; altered/evaluable patients shown above bars. Adjusted OR for ATRX/DAXX loss in uterine versus extrauterine tumors, 1.96 (95% CI, 1.49 to 2.58; P* < .001)

**Fig 4.**
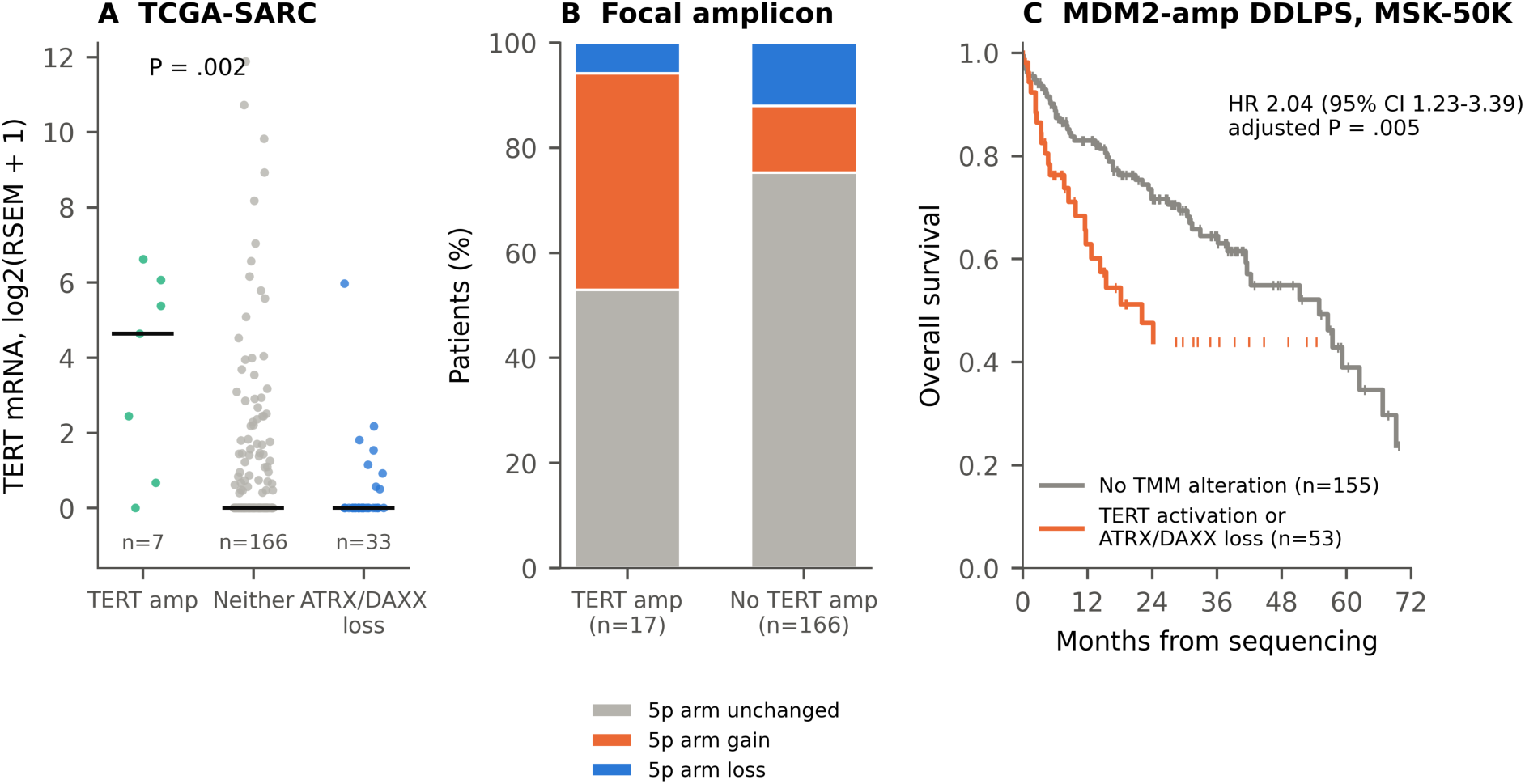
Validation of the liposarcoma findings. (A) TERT mRNA expression in TCGA-SARC by telomere maintenance status; horizontal bars, medians (Mann-Whitney P, TERT amplification v neither). (B) Arm-level 5p status in MDM2-amplified DDLPS from MSK-IMPACT 50K with and without TERT amplification. (C) Overall survival from sequencing in MDM2*-amplified DDLPS (MSK-IMPACT 50K) with versus without a telomere maintenance alteration; tick marks denote censoring. HR adjusted for age, sample type, and metastatic disease status at sequencing

### Germline Telomere Length and Sarcoma Susceptibility

Because telomere maintenance was acquired during progression from WDLPS to DDLPS, we used FinnGen to test whether germline telomere length is associated with liposarcoma risk.

Of 124 telomere length instruments, 101 were harmonized with FinnGen (mean F statistic, 134). Genetically predicted longer telomere length was associated with liposarcoma incidence (IVW OR per SD, 4.10; 95% CI, 2.15 to 7.81; *P* = 1.8 × 10^−5^; false discovery rate *q* < .001; Fig 5A). The estimate was consistent with the weighted median (OR, 3.47; 95% CI, 1.27 to 9.46) and MR-Egger (OR, 3.27; 95% CI, 1.09 to 9.84), without evidence of directional pleiotropy (Egger intercept *P* = .62) or heterogeneity (Cochran *Q P* = .32), and persisted after exclusion of palindromic variants (OR, 4.06; 95% CI, 1.99 to 8.29) and of canonical telomere-biology loci (OR, 3.08; 95% CI, 1.50 to 6.32; Fig 5B-C). No single variant drove the association (leave-one-out ORs, 3.10 to 4.51; all *P* < .001).

**Fig 5.**
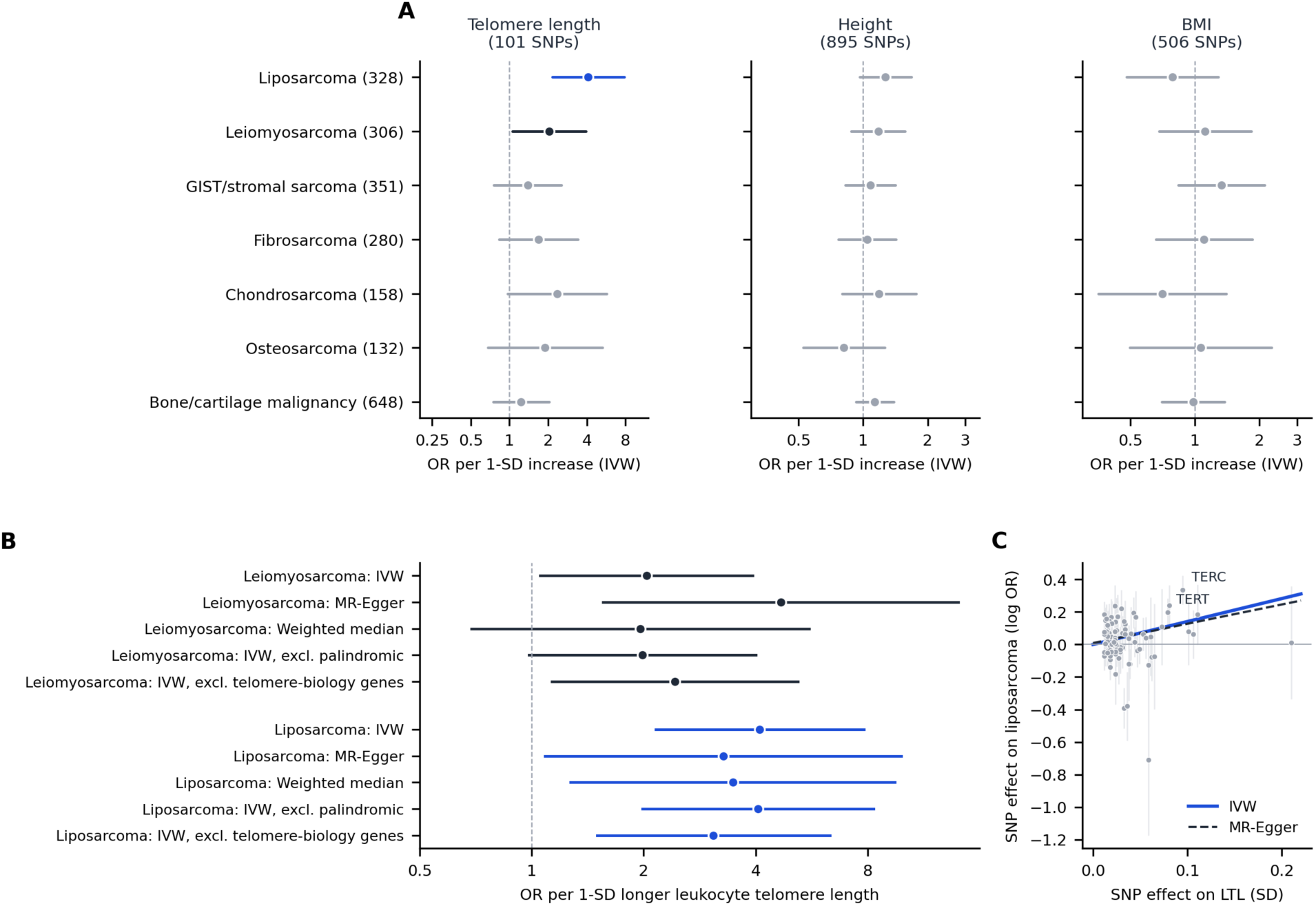
Two-sample Mendelian randomization of leukocyte telomere length, height, and body mass index on sarcoma risk in FinnGen release 12. (A) Inverse-variance weighted ORs per SD increase in each exposure; case numbers in parentheses; blue, false discovery rate < 0.05; black, nominal P < .05. (B) Sensitivity analyses for telomere length with liposarcoma and leiomyosarcoma; telomere-biology genes include TERT, TERC, and PARP1. (C) Variant-level effects on telomere length and liposarcoma with IVW and MR-Egger slopes; error bars, SE; variants with outcome SE above 0.5 are omitted for display.

Longer telomere length was nominally associated with leiomyosarcoma (OR, 2.03; 95% CI, 1.05 to 3.93; *P* = .034; *q* = .36), though this did not survive correction for multiple testing. Neither height nor body mass index was associated with any endpoint after correction for multiple testing. With 132 to 648 cases per endpoint, the minimum OR detectable at 80% power ranged from 2.0 to 4.3 per SD of telomere length; the absence of associations for other subtypes therefore does not exclude effects.

## DISCUSSION

In this analysis of 11,792 sequenced sarcomas, telomerase activation and ALT rarely co-occurred and segregated by histology. First, a substantial fraction of telomerase activation in sarcoma proceeds through *TERT* amplification and rearrangement rather than promoter mutation, and these events account for most of the observed mutual exclusivity. Second, DDLPS activates telomerase through *TERT* amplification or rearrangement, never through promoter mutation, in a pattern strictly exclusive of ATRX/DAXX loss. Third, telomere maintenance alterations are acquired at liposarcoma dedifferentiation, rising from 8.7% in WDLPS to 28.0% in DDLPS. Fourth, telomere length was associated with risk of liposarcoma by Mendelian analysis.

Previous studies characterized DDLPS primarily as an ALT-driven tumor.^11^ Our data indicate that DDLPS comprises two mechanistically distinct telomere maintenance subsets of comparable size: approximately 20% of the tumors activate telomerase through *TERT* copy-number or structural alteration, and approximately 10% use ALT. The complete absence of promoter mutations in *MDM2*-amplified DDLPS contrasts sharply with myxoid/round cell liposarcoma, in which promoter mutation is nearly universal.^8,18^ This contrast suggests that the genomic route to telomerase reactivation is constrained by the dominant oncogenic program. The 12q-amplified, genomically unstable DDLPS genome, in which scattered amplification is a recurring feature,^19^ may favor *TERT* activation, whereas the FUS::DDIT3-driven, copy-number-quiet myxoid genome instead acquires a point mutation.

The strict exclusivity of *TERT* amplification with ATRX/DAXX loss supports functional redundancy. Independence from *CDK4* and *JUN* status, *TERT* transcription in amplified tumors, and a focal amplicon leaving the 5p arm intact in most cases all support this interpretation.

The enrichment of both mechanisms in DDLPS relative to WDLPS suggests that telomere maintenance may enable dedifferentiation to evolve. WDLPS is indolent and genomically simpler, whereas DDLPS acquires additional copy-number complexity, including 1p32 and 6q23 amplification, and aggressive behavior.^12,13^ Telomere dysfunction preceding telomere maintenance is a recognized driver of chromosomal instability.^20^ Paired analysis of well-differentiated and dedifferentiated components within the same tumor would be helpful to confirm. If confirmed, *TERT* amplification or ATRX/DAXX loss in an atypical lipomatous tumor or WDLPS could serve as a biomarker of dedifferentiation risk.

DDLPS with telomerase activation or ALT had a median survival less than half that of DDLPS without either alteration (54.9 vs 22.1 months). Current DDLPS risk assessment relies on grade, site, and resection status. Our data suggest that *TERT* activation or ALT may add prognostic information beyond these variables and warrants prospective testing.

Our data suggests a mechanism-matched framework for clinical trial design in DDLPS (Table 3). Systemic options for advanced DDLPS remain limited, and MDM2 and CDK4 inhibitors and immune checkpoint blockade have produced modest activity in unselected populations.^13^ Telomere maintenance status, already captured by routine panel sequencing, defines two mutually exclusive subsets that together account for approximately 28% of *MDM2*-amplified DDLPS and could be prospectively enriched. Patients with liposarcoma with the tumor that carries *TERT* amplification or rearrangement could be candidates for telomerase-directed therapy. Patients with liposarcoma with the tumors with ATRX/DAXX loss could be candidates for agents that exploit replication stress at ALT telomeres, including ATR inhibitors, to which ALT-positive cells were hypersensitive in preclinical models.^21^

**Table 3.**
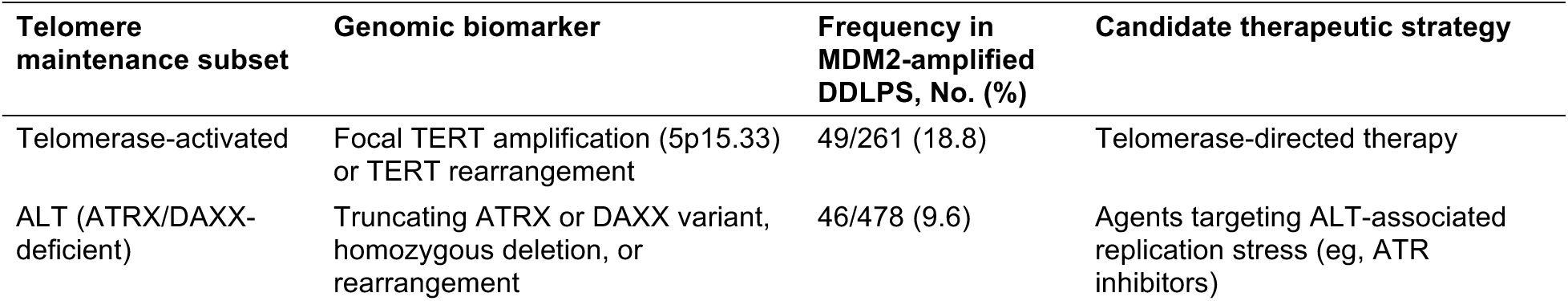
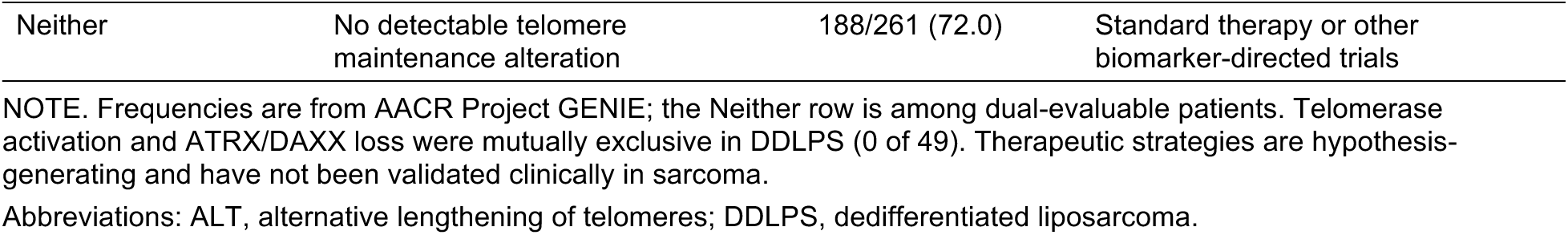
Proposed Mechanism-Matched Stratification of Dedifferentiated Liposarcoma by Telomere Maintenance Status.

The enrichment of ATRX/DAXX loss in uterine leiomyosarcoma supports the previous reports of frequent *ATRX* alteration in uterine primaries^22^ and ALT in leiomyosarcoma broadly.^23^ The site difference persisted after adjustment for age and center and was not explained by sex. Whether it reflects distinct cells of origin or differences in the smooth muscle differentiation program between uterine and extrauterine leiomyosarcoma merits investigation with transcriptomic and epigenomic data.

Genetically longer telomeres were associated with approximately fourfold higher odds of liposarcoma per SD, supporting the evidence that longer telomeres increase the risk of several cancers, particularly those arising in tissues with low rates of stem cell division,^34^ and with the enrichment of heritable defects in telomere maintenance genes among patients with sarcoma.^7^ A longer replicative reserve may allow an adipocytic precursor with 12q13-15 amplification to accumulate additional alterations before reaching telomere crisis, with somatic *TERT* amplification or ALT at dedifferentiation marking the escape from that crisis. Because FinnGen does not resolve liposarcoma subtypes, the germline association cannot be assigned to WDLPS or DDLPS specifically, and replication in a subtype-resolved cohort is required.

Our study has limitations. First, ATRX/DAXX inactivation is a genomic proxy for ALT; tumors with ALT but no detectable ATRX/DAXX alteration were misclassified as negative, and the ALT phenotype was not confirmed by C-circle assay or telomere-specific fluorescence in situ hybridization. Second, copy-number calls in GENIE were discrete panel-based estimates; expression and focality were confirmed in validation cohorts of modest size, and amplitude could not be quantified reliably. Third, GENIE aggregates heterogeneous assays; although we restricted denominators to evaluable patients and adjusted for center, residual assay effects cannot be excluded, and promoter evaluability excluded roughly half of the cohort. Fourth, WDLPS and DDLPS were sampled from different patients. Finally, outcome analyses relied on a single institution overlapping the discovery cohort, and lacked grade and treatment data; these results should be considered hypothesis-generating until replicated in an independent, clinically annotated cohort.

In conclusion, telomere maintenance in sarcoma is histology-specific, and its two mechanisms rarely co-occur. In liposarcoma, telomere maintenance is acquired at dedifferentiation through focal *TERT* amplification or ALT rather than promoter mutation, and it identifies DDLPS with inferior survival. Germline genetic evidence further implicates telomere length in liposarcoma susceptibility.

## Data Availability

All data analyzed in this study are publicly available: AACR Project GENIE (v20.0-public) and cBioPortal (TCGA-SARC, MSK-IMPACT 50K, MSK-MET, and the Foundation Medicine sarcoma cohort); FinnGen release 12 summary statistics; and published genome-wide association summary statistics for leukocyte telomere length (UK Biobank; Codd et al., Nat Genet 2021) and for height and body mass index (GIANT-UK Biobank; Yengo et al., Hum Mol Genet 2018). Analysis code is available from the corresponding author on reasonable request.

https://genie.cbioportal.org

https://www.cbioportal.org/

https://www.finngen.fi/en/access_results

## Author contributions

MP, concept, data analysis and writing manuscript and revision; JC, data analysis; LZ, data analysis; MZ, data analysis and revision, NB, data analysis and revision; KG, revision.

## Acknowledgment

The authors acknowledge the American Association for Cancer Research and its financial and material support in the development of the AACR Project GENIE registry, as well as members of the consortium for their commitment to data sharing. The authors also acknowledge the participants and investigators of the FinnGen study.

## Data Availability

All analyses derive from public cBioPortal datasets, FinnGen release 12 summary statistics, and published genome-wide association summary statistics.

## Data sources

AACR Project GENIE v20.0-public; TCGA-SARC; MSK-IMPACT 50K; MSK-MET; Foundation Medicine sarcoma cohort; FinnGen release 12; published genome-wide association studies of leukocyte telomere length, height, and body mass index (all accessed September 2026)

## Declaration of generative AI in scientific writing

Claude (Opus 5.5; Anthropic, San Francisco, CA) and Gemini 3.0 were used to assist with data retrieval through the cBioPortal application programming interface, development of Python analysis code, statistical analysis, figure generation, validation and literature search. The authors conceived the study, independently reviewed and verified the analyses, code, data sources, and cited references, edited the text, and take full responsibility for the accuracy and integrity of the data and conclusions.

